# Effect of Early Treatment with Fluvoxamine on Risk of Emergency Care and Hospitalization Among Patients with COVID-19: The TOGETHER Randomized Platform Clinical Trial

**DOI:** 10.1101/2021.08.19.21262323

**Authors:** Gilmar Reis, Eduardo Augusto dos Santos Moreira Silva, Daniela Carla Medeiros Silva, Lehana Thabane, Aline Cruz Milagres, Thiago Santiago Ferreira, Castilho Vitor Quirino dos Santos, Adhemar Dias de Figueiredo Neto, Eduardo Diniz Callegari, Leonardo Cançado Monteiro Savassi, Vitoria Helena de Souza Campos, Ana Maria Ribeiro Nogueira, Ana Paula Figueiredo Guimaraes Almeida, Maria Izabel Campos Simplicio, Luciene Barra Ribeiro, Rosemary Oliveira, Ofir Harari, Jamie I Forrest, Hinda Ruton, Sheila Sprague, Paula McKay, Alla V Glushchenko, Craig R. Rayner, Eric J. Lenze, Angela M. Reiersen, Gordon H. Guyatt, Edward J. Mills, for the TOGETHER Investigators

## Abstract

**Background:** Recent evidence indicates a potential therapeutic role of fluvoxamine for COVID-19. In the TOGETHER randomized platform clinical trial for acutely symptomatic patients with COVID-19, we assessed the efficacy of fluvoxamine vs. placebo in preventing either extended emergency room observation or hospitalization due to COVID-19. Herein, we report the preliminary findings.

**Methods:** This placebo-controlled, randomized, adaptive, platform trial conducted among symptomatic Brazilian adults confirmed positive for SARS-CoV-2 included eligible patients with a known risk factor for progression to severe disease. Patients were randomly assigned to either fluvoxamine (100 mg twice daily for 10 days) or placebo. The primary endpoint was a composite outcome of emergency room observation for >6 hours or hospitalization from COVID-19 up to 28 days post randomization using intention to treat. Modified intention to treat (mITT) explored patients receiving at least 24 hours of treatment before a primary outcome event. Secondary outcomes included viral clearance at day 7, time to hospitalization, mortality, and adverse drug reactions. We used a Bayesian analytic framework to determine effects along with probability of success of intervention compared to placebo. The trial is registered at clinicaltrials.gov (NCT04727424) and is ongoing.

**Findings:** The study team screened 9020 potential participants for this trial. The trial was initiated on June 2, 2020, with the current protocol reporting randomization from January 15, 2021 to August 6^th^ 2021, when the trial arms were stopped for superiority. A total of 3238 patients were allocated to fluvoxamine (n=739), placebo (n=733) and other treatments (n=1766). Herein, we report the effectiveness of fluvoxamine vs. a concurrent placebo control. The average age of participants was 50 years (range 18-102 years); 57% were female. The proportion of patients observed in an emergency room for >6 hours or admitted to hospital due to COVID-19 was lower for the fluvoxamine group compared to placebo (77/739 vs 108/733; Relative Risk [RR]: 0.71; 95% Bayesian Credible Interval [95% BCI]: 0.54 - 0.93), with a probability of superiority of 99.4% surpassing the prespecified superiority threshold of 97.6% (risk difference 4.3%). Of the composite primary outcome events, 88% were hospitalizations. Findings were similar for the mITT analysis (RR0.68, 95% BCI : 0.50- 0.91). We found no significant relative effects between the fluvoxamine and placebo groups on viral clearance at day 7 (Odds ratio [OR]: 0.75; 95% Confidence Intervals [95% CI]: 0.53 - 1.07), mortality (OR: 0.70; 95% CI: 0.36 - 1.30), time to death (Hazard ratio [HR]: 0.79; 95% CI: 0.58 - 1.08), days hospitalized (Mean Difference (MD) 1.22 days; 95% CI: 0.98 - 1.53), number of days ventilated (MD 1.10; 95% CI: 0.70 - 1.73) or other secondary outcomes. Data capturing all 28 days of follow-up will be reported after August 26^th^, 2021.

**Interpretation:** Treatment with fluvoxamine (100 mg twice daily for 10 days) among high-risk outpatients with early diagnosed COVID-19, reduced the need for extended emergency room observation or hospitalization.

**Funding:** The trial was supported by FastGrants and The Rainwater Foundation.

## BACKGROUND

Although safe and effective vaccines for COVID-19 have been developed and distributed, there remain, particularly in low resource settings, major challenges regarding their production, allocation, and affordability.^3^ Identifying inexpensive, widely available and effective therapies against COVID-19 is, therefore, of great importance. In particular, repurposing existing medicines that are widely available and with well understood safety profiles, has particular appeal.^4^

Fluvoxamine is a selective serotonin reuptake inhibitor (SSRI) and a Sigma-1 receptor (S1R) agonist.^5^ There are several potential mechanisms for fluvoxamine in treatment of COVID-19 illness, including anti-inflammatory and possible antiviral effects.^6^ A small placebo-controlled randomized trial has raised the possibility that fluvoxamine may reduce the risk of clinical deterioration in outpatients with COVID-19, suggesting the need for larger randomized, placebo-controlled studies.^1,2^

To evaluate the efficacy of fluvoxamine to prevent progression of COVID-19 and hospitalization among outpatients with laboratory-documented SARS-CoV-2, we conducted a randomized, placebo-controlled adaptive platform trial in Minas Gerais, Brazil. This flexible platform trial design allows for additional agents to be added and tested with standardized operating procedures outlined in a single overarching master protocol.^7,8^ Among eight different interventions evaluated in this platform trial, we report here on the clinical evaluation of fluvoxamine using a concurrent placebo control group.

## METHODS

### Study design and oversight

The TOGETHER Trial is a randomized adaptive platform trial to investigate the efficacy of repurposed treatments for COVID-19 disease among high-risk adult outpatients.^9^ The trial was designed and conducted in partnership with local public health authorities from eleven participating cities in Brazil to simultaneously test potential treatments for early disease using a master protocol. A master protocol defines prospective decision criteria for discontinuing interventions for futility, stopping due to superiority against placebo, or adding new interventions. Interventions evaluated in the TOGETHER trial, thus far, include, hydroxychloroquine (protocol 1), lopinavir/ritonavir (protocol 1),^10^ metformin, ivermectin, fluvoxamine, doxasozin and pegylated interferon lambda versus matching placebos (protocol 2).

The trial began on June 2, 2020 and enrollment into the fluvoxamine arm began on January 15^th^ 2021. The trial, which complied with the International Conference of Harmonization – Good Clinical Practices, as well as local regulatory requirements, was approved for research ethics by local and national ethics boards in Brazil (CONEP CAAE: 41174620.0.1001.5120, approval letter 5.501.284) and the Hamilton Integrated Research Ethics Board (HiREB, approval letter 13390) in Canada. The full protocol, statistical analysis plan, and additional details are appended in the web-appendix. The adaptive designs CONSORT extension (ACE) statement guided this trial report.^16,17^ The steering committee made all protocol-related decisions and sponsors had no role in trial conduct, data analysis or decision to submit manuscript for publication. An independent Data Safety Monitoring Committee (DSMC) provided trial oversight.

## Setting

The appended web-appendix lists the cities and investigators of the eleven clinical sites in Brazil that participated in the trial. Local investigators, in partnership with local public health authorities, recruited participants at community health facilities (emergency settings, flu-symptom referral centers, primary care community centers). We used several community outreach strategies including physical and social media as per local public health authorities, in order to create awareness of the trial.

### Participant Screening

Upon presentation to one of the trial outpatient care clinics, local investigators screened potential participants to identify those who met the eligibility criteria.

The key inclusion criteria were: 1) patients over the age of 18, 2) presenting to an outpatient care setting with an acute clinical condition consistent with COVID-19 and symptoms beginning within 7 days of the screening date, 3) positive rapid test for SARS-CoV-2 antigen performed at the time of screening or patient with positive SARS-CoV-2 diagnostic test within 7 days of symptom onset, 4) at least one additional criterion for high-risk: diabetes mellitus; systemic arterial hypertension requiring at least one oral medication for treatment; known cardiovascular diseases (heart failure, congenital heart disease, valve disease, coronary artery disease, cardiomyopathies being treated, clinically manifested heart disease and with clinical repercussion); symptomatic lung disease and / or being treated (emphysema, fibrosing diseases); symptomatic asthma patients requiring chronic use of agents to control symptoms; smoking; obesity, defined as BMI>30 kg / m^2^ (weight and height information provided by the patient); transplant patients; patient with stage IV chronic kidney disease or on dialysis; immunosuppressed patients / using corticosteroid therapy (equivalent to at least 10 mg of prednisone per day) and / or immunosuppressive therapy; patients with a history of cancer in the last 0.5 years or undergoing current cancer treatment, or age ≥ 50 years; and unvaccinated status.

Patients who met any of the following criteria were excluded from the trial: 1) Diagnostic examination for SARS-CoV2 negative associated with acute flu-like symptoms (patient with negative test taken early and becoming positive a few days later were eligible, if he/she was <7 days after the onset of flu-like symptoms); 2) Acute respiratory condition compatible with COVID-19 treated in the primary care and previously requiring hospitalization; 3) Acute respiratory condition due to other causes; 4) Received vaccination for SARS-CoV2; 5) Dyspnea secondary to other acute and chronic respiratory causes or infections (e.g., decompensated COPD, acute bronchitis, pneumonia, primary pulmonary arterial hypertension); 6) Current use of selective serotonin reuptake inhibitors; uncontrolled psychiatric disorders; or suicidal ideation; 7) Inability or unwillingness to follow research guidelines and procedures. A full list of exclusion criteria are provided in the trial protocol.

If a patient met the above eligibility criteria, study personnel obtained written informed consent. After obtaining informed consent a rapid antigen test for COVID-19 (Panbio ®, Abbott laboratories) and a pregnancy test for women of childbearing age were performed. If the COVID-19 test was negative or if the pregnancy test was positive, the participant was not included in the trial. After informed consent, study personnel collected the following data prior to randomization: demographics, medical history concomitant medications, co-morbidities, exposure to Index Case information, WHO clinical worsening scale, and the PROMIS Global Health Scale.

### Randomization and Trial Interventions

Participants were randomized using a centralized core randomization process handled by an independent unblinded pharmacist who was not aware of any protocol-related procedures and contracted specifically for this process. Sites requested randomization by text message to the pharmacist at the coordinating center. This maintained concealment of allocation. Patients were randomly assigned using a block randomization procedure for each participating site, stratified by age (< 50 years /≥ 50 years). The trial team, site staff and patients were blinded to treatment allocation. The active drugs and the placebo pills were packaged in identically shaped bottles and labeled with alphabet letters corresponding to the active arm or placebo arm. Only the third-party pharmacist responsible for releasing the randomization was aware of which letter was associated with which drug and/or placebo. As this is a multi-arm trial and all active interventions have a matching inert placebo, the matching placebo represents the proportion of the control group for the number of arms in the trial at any given time.

### Data Collection and Participant Follow-Up

Our primary outcome is a composite that includes emergency room visits due to the clinical worsening of COVID-19 (defined as participant remaining under observation for > 6 hours) or hospitalization due to the progression of COVID-19 (worsening of clinical status) and/or suspected COVID-19 complications within 28 days of randomization. Key secondary outcomes include: 1) viral clearance, 2) time to clinical improvement, 3) number of days with respiratory symptoms, 4) time to hospitalization for any cause or due to COVID-19 progression, 5) all-cause mortality and time to death from any causes, 7) WHO clinical worsening scale score, 8) days in hospital and on ventilator and 9) adverse events, adverse reactions to the study medications and the proportion of participants who are non-adherent with the study drugs. All secondary outcomes are assessed up to 28 days following randomization.

Study personnel collected, via in person, telephone contact and social media applications using video-teleconferencing, outcome data on days 1, 2, 3, 4, 5, 7, 10, 14, and 28. We collected outcome data irrespective of whether participants took study medication. In case of adverse events, unscheduled visits (during the treatment period) outside of clinical care could occur at any time.

Considering the transmissible characteristic of SARS-CoV-2 and the isolation recommendations of positive individuals, we collected limited vital sign data. Cardiac safety was assessed using a 6-lead ECG (Kardiamobile, Mountain View, CA) at the baseline visit. The digital recordings were de-identified and transferred to a central facility (Cardresearch, Belo Horizonte, Brazil) for reading. Oxygen status was assessed using a pulse oximeter for non-invasive arterial oxygen saturation (SpO2) and pulse (Jumper Medical Equipment, Shenzhen, China), and temperature using a standard digital oral thermometer administered by research personnel. Mid-turbinate nasal swab kits and sterile recipient storage were provided for collection of nasopharyngeal swab or sputum/saliva. They were performed by the first quarter of participants enrolled in the trial at days 3 and 7. PCR of viral clearance was assessed to determine if active drugs demonstrate any anti-viral effects.

All serious and non-serious adverse events were reported to study personnel as per local regulatory requirements. Reportable adverse events included serious adverse events, adverse events resulting in study medication discontinuation, and adverse events assessed as possibly related to study medication.

### Trial interventions

All participants received usual standard care for COVID-19 provided by healthcare professionals workers at public health facilities. Patients were randomized to fluvoxamine (Luvox ®, Abbott) at a dose of 100 mg twice a day for 10 days or corresponding placebo starting right after randomization (day 1). Research personnel provided participants a welcome video with information on trial, study drug, adverse events and follow-up procedures. Clinicians providing usual care in public health facilities typically focus on the management of symptoms and with antipyretics, and recommend antibiotics only if they suspect bacterial pneumonia.

### Statistical Analyses

The Adaptive Design Protocol and the Master Statistical Analysis Plan provide details of sample size calculation and statistical analysis (appended). This trial is adaptive and applies sample size re-estimation approaches. To plan for each arm, we assumed a minimum clinical utility of 37.5% (relative risk reduction) to achieve 80% power with 0.05 two-sided Type 1 error for a pairwise comparison against the placebo (talc) assuming a control event rate (CER) of 15%. This resulted in an initial plan to recruit 681 participants per arm. The statistical team conducted planned interim analyses. Stopping thresholds for futility were established if the posterior probability of superiority was less than 40% at interim analysis. An arm can be stopped for superiority if the posterior probability of superiority meets the threshold of 97.6%

Baseline characteristics are reported as count (percent) or median and interquartile range (IQR)for continuous variables. We applied a Bayesian framework for our primary outcome analysis and a frequentist approach for all sensitivity analyses and secondary outcomes. Posterior efficacy of fluvoxamine for the primary outcome is calculated using the beta-binomial model for event rates, assuming informed priors based on the observational data for both placebo and fluvoxamine, for both Intention-to-Treat (ITT), modified ITT (defined as receiving the study drug for at least 24 hours before an event) and Per-Protocol (PP) analyses (defined as taking >80% of possible doses). Modified ITT was defined as receiving treatment for at least 24 hours before a primary outcome. We accounted for any temporal changes in events rates by using only the concurrent randomised population. We assessed subgroup effects according to the preplanned statistical analysis plan. We calculated the number needed to treat.

Secondary outcomes were assessed using a pre-specified frequentist approach. For viral clearance we fitted a longitudinal, mixed-effect logistic regression model with a treatment and time interaction term for binary patient outcomes (Covid-19 positive/negative) reported on day 3 and 7 from randomization, with subject random effect. We assessed time-to-event outcomes using Cox proportional hazard models and binary outcomes using logistic regression. Per protocol analyses were considered sensitivity analyses to assess the robustness of the results. All analyses were performed using R version 4.0.3. Full details of the Statistical Analysis Plan (SAP) are appended.

### Data and Safety Monitoring Committee

A Data and Safety Monitoring Committee provided independent oversight for this trial. We planned a 4^th^ interim analysis of the fluvoxamine arm with data collected up to August 2^nd^, 2021.

## RESULTS

We screened 9020 potential participants for inclusion in this trial to date. The trial enrolled its first participant on June 2, 2020 and enrolment into the fluvoxamine arm began on January 20, 2021. By August 6, 2021, 1,472 recruited participants were randomized to fluvoxamine (n= 739) or the placebo (n=733), and 1,766 were randomized to other treatment arms (Figure 1). The median age was 50 years (range 18-102) and 846 (57.5%) were women (Table 1). Most participants self-identified as mixed-race 1,403 (95.3%), 11 (0.7%) as white, 10 (0.7%) as black or African American, the rest self-identified as unknown 48 (3.3%). As the trial is ongoing, herein we provide descriptive summaries of only those randomized to fluvoxamine and its concurrent control. With respect to covariates of age, BMI, and co-morbidities, the groups were generally well balanced (Table 1). The mean number of days with symptoms prior to randomization was 4 days (Standard Deviation 1.76).

**Table 1.** Patient characteristics by treatment allocation in the TOGETHER Trial

|  | **Fluvoxamine**<br>(n=739) | **Placebo**<br>(n=733) | **Total**<br>(n=1472) |
| --- | --- | --- | --- |
| **Sex** |  |  |  |
| Female | 408(55.2) | 438(59.8) | 846(57.5) |
| Male | 331(44.8) | 295(40.2) | 626(42.5) |
| **Race** |  |  |  |
| Mixed Race<sup>+</sup> | 708(95.8) | 695(94.8) | 1403(95.3) |
| White | 5(0.7) | 6(0.8) | 11(0.7) |
| Black or African American | 5(0.7) | 5(0.7) | 10(0.7) |
| Unknown | 21(2.8) | 27(3.7) | 48(3.3) |
| **Age, years** |  |  |  |
| >= 50 years | 325(44.0) | 318(43.4) | 643(43.7) |
| **Age Descriptive Statistics** |  |  |  |
| Median | 50 | 49 | 50 |
| IQR | 17 | 18 | 18 |
| **Body Mass Index (BMI)** |  |  |  |
| <30 kg/m<sup>2</sup> | 354(47.9) | 360(49.1) | 714(48.5) |
| >=30 kg/m<sup>2</sup> | 372(50.3) | 359(49.0) | 731(49.7) |
| Unspecified | 13(1.8) | 14(1.9) | 27(1.8) |
| **Time since onset of symptoms** |  |  |  |
| 0-3 days | 318(43.0) | 296(40.4) | 614(41.7) |
| 4-7 days | 241(32.6) | 249(34.0) | 490(33.3) |
| Unspecified | 180(24.3) | 188(25.7) | 368(25.0) |
| **Risk factors** |  |  |  |
| Chronic cardiac disease | 8(1.1) | 8(1.1) | 16(1.1) |
| Hypertension | 106(14.4) | 88(12.0) | 194(13.2) |
| Chronic pulmonary disease | 6(0.8) | 3(0.4) | 9(0.6) |
| Asthma | 11(1.5) | 16(2.2) | 27(1.8) |
| Chronic kidney disease | 2(0.3) | 2(0.3) | 4(0.3) |
| Rheumatologic disorder | 1(0.1) | 0(0.0) | 1(0.1) |
| Chronic neurological disorder | 8(1.1) | 6(0.8) | 14(1.0) |
| Diabetes mellitus: Type 1 | 25(3.4) | 22(3.0) | 47(3.2) |
| Diabetes mellitus: Type 2 | 103(14.0) | 93(12.7) | 196(13.3) |
| Obesity | 2(0.3) | 1(0.1) | 3(0.2) |
| Any other risk factor(s) or co-morbidities | 24(3.3) | 19(2.6) | 43(2.9) |

**LEGEND:** <sup>†</sup>Self-identified as someone with mixed-race ancestry.

**Figure 1:**
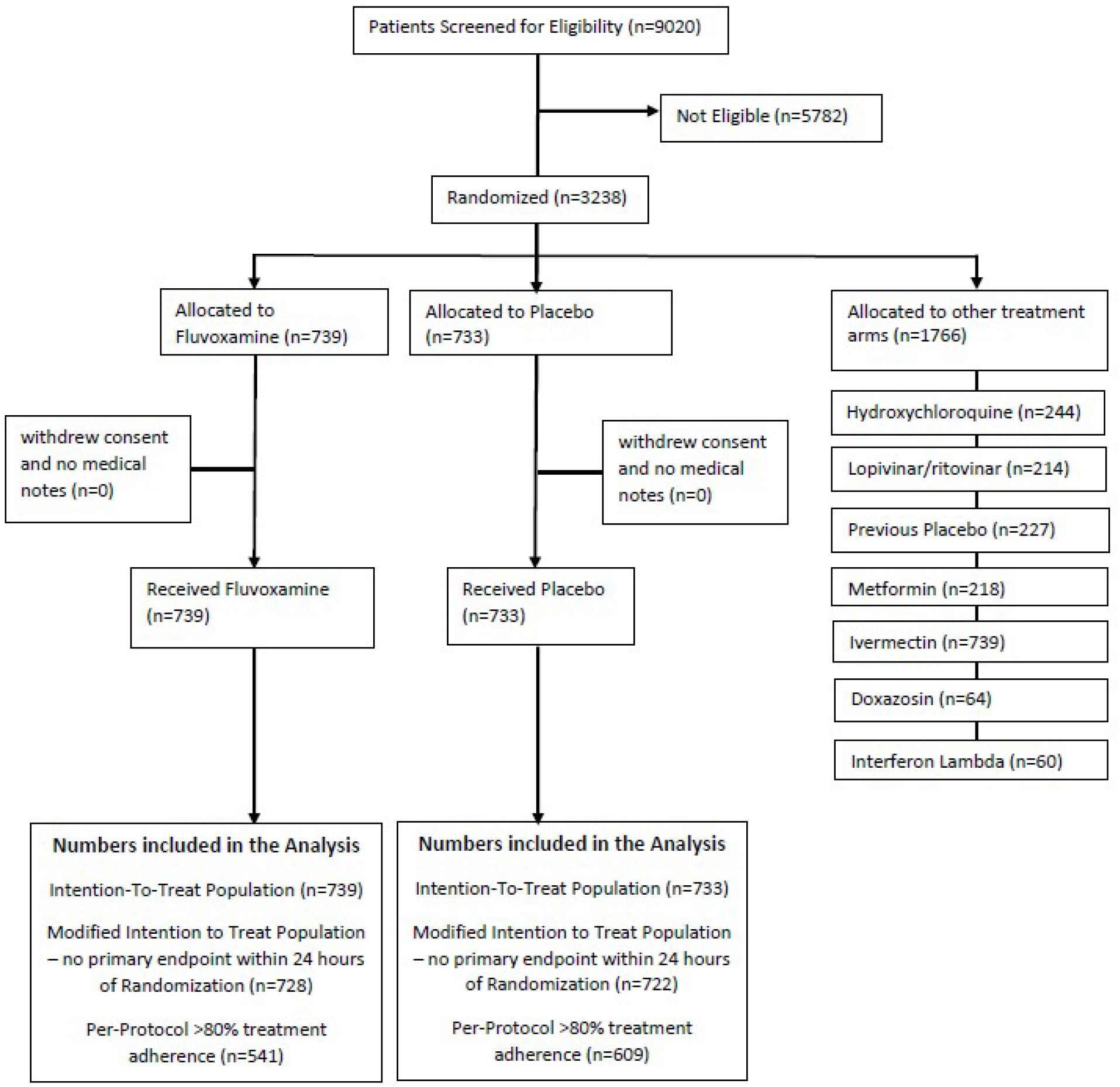
Participant Flowchart.

### Primary Outcomes

The Data Safety Monitoring Committee met four times since the protocol initiation and last met on August 5, 2021, recommending that the TOGETHER Trial stop randomizing patients to the fluvoxamine arm, as this comparison had met the pre-specified superiority criterion for the primary endpoint (prespecified superiority threshold 97.6%). Based on the Bayesian beta-binomial model, there was evidence of a benefit of fluvoxamine reducing hospitalization or observation in an emergency room for greater than six hours due to COVID-19 (Relative Risk [RR]: 0.71; 95% Bayesian Credible Interval [BCI]: 0.54 - 0.93) in the Intention-to-Treat (ITT) population (figure 2A) and (RR: 0.68; 95% BCI: 0.50 - 0.91) in a modified ITT population (figure 2B). The probability that the event rate was lower in the fluvoxamine group compared to placebo was 99.4% for the ITT population, and 99.6% for the modified ITT population (Figure 2A/B). These posterior efficacy numbers were higher than the pre-specified 97.6% threshold set for the fourth interim analysis. In the fluvoxamine group 77 (10.4%) participants experienced a primary outcome event compared to 108 (14.7%) in the placebo group (Table 2). Most events (88%) were hospitalizations. The number needed to treat is 24. Per protocol analysis demonstrated a larger treatment effect (0.34, 95% BCI, 0.20-0.54).

**Table 2:** Proportion of primary outcome events and relative risk of extended emergency room observation or hospitalization of fluvoxamine vs. placebo

|  | Intention-to-treat analysis |  |  | Modified intention-to-treat analysis |  |  |
| --- | --- | --- | --- | --- | --- | --- |
|  | N | n (%) | Relative risk<br>(95% CrI) | N | n (%) | Relative risk<br>(95% CrI) |
| Fluvoxamine | 739 | 77 (10.4 %) | 0.71 [0.54; 0.93] | 728 | 66(9.1 %) | 0.68 [0.50; 0.91] |
| Placebo | 733 | 108 (14.7 %) | 1.00 (ref) | 722 | 97(13.4 %) | 1.00 (ref) |
| All | 1472 | 185 (12.6%) | -- | 1450 | 163(11.2 %) | -- |

**LEGEND:** 95% CrI – Credible intervals

**Figure 2:**
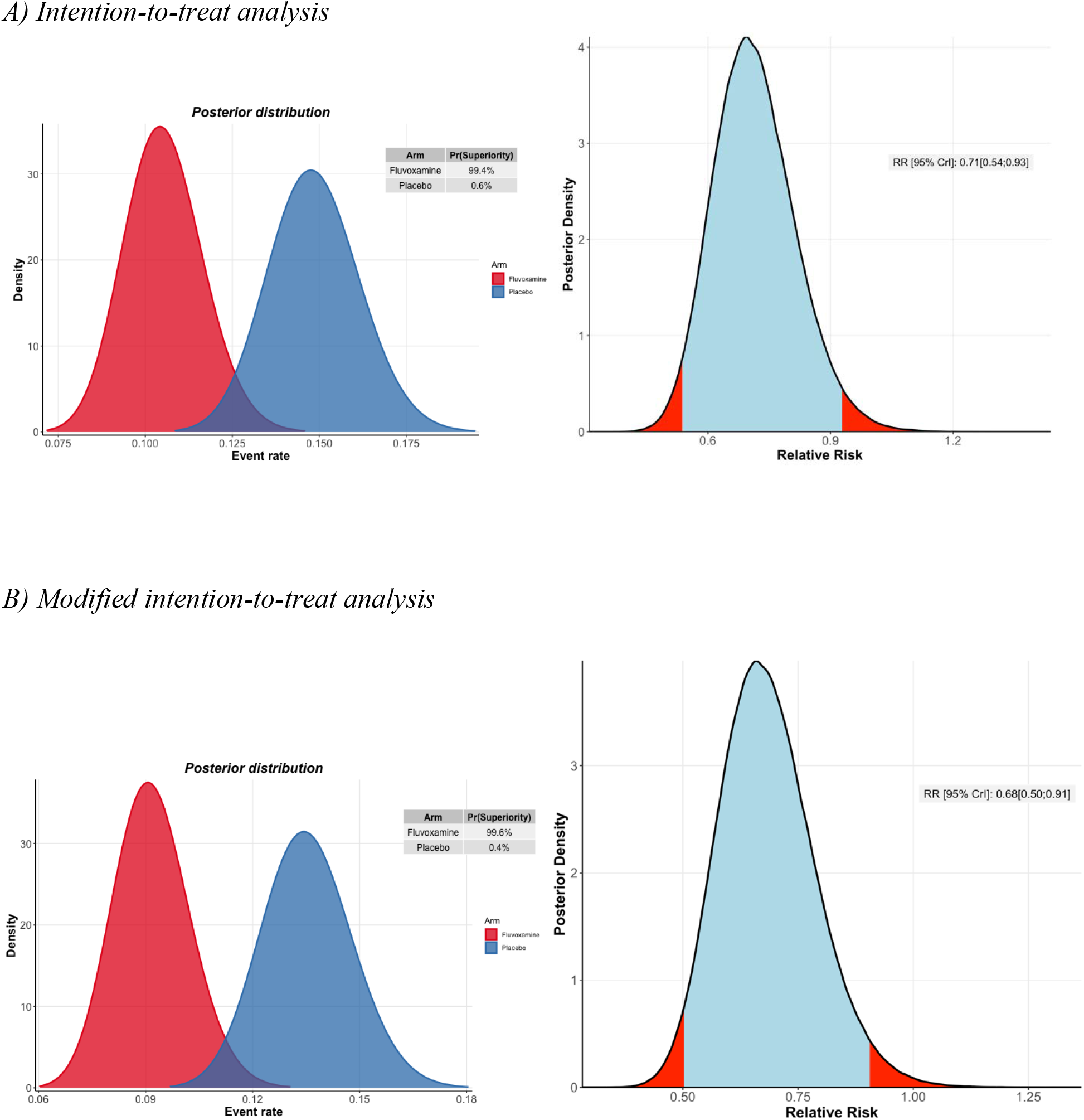
Probability of efficacy and Bayesian relative risk of extended emergency room observation or hospitalization for fluvoxamine vs. placebo (Panel A: ITT population; Panel B: Modified ITT population)

### Secondary Outcomes

Table 3 presents findings from secondary outcome analyses. There were no significant differences between fluvoxamine and placebo for viral clearance at Day 7 (p=0.18) and eFigure3 in web-appendix, hospitalizations due to COVID (p=0.17), all-cause hospitalizations (p=0.12), time to hospitalization (p=0.14, number of days in hospital (p=0.07), mortality (p=0.26), time to death (p=0.26), number of days on mechanical ventilation (p=0.67), time to recovery (p=0.86) or the PROMIS Global Health Scale (p=0.44). With respect to adverse events, there were significantly greater number of Grade 1 (mild) treatment emergent adverse events (TEAE) among patients in the fluvoxamine arm (p<0.01). However, no differences between fluvoxamine and placebo were observed for TEAEs of Grades 2, 3, 4, or 5.

**Table 3:**
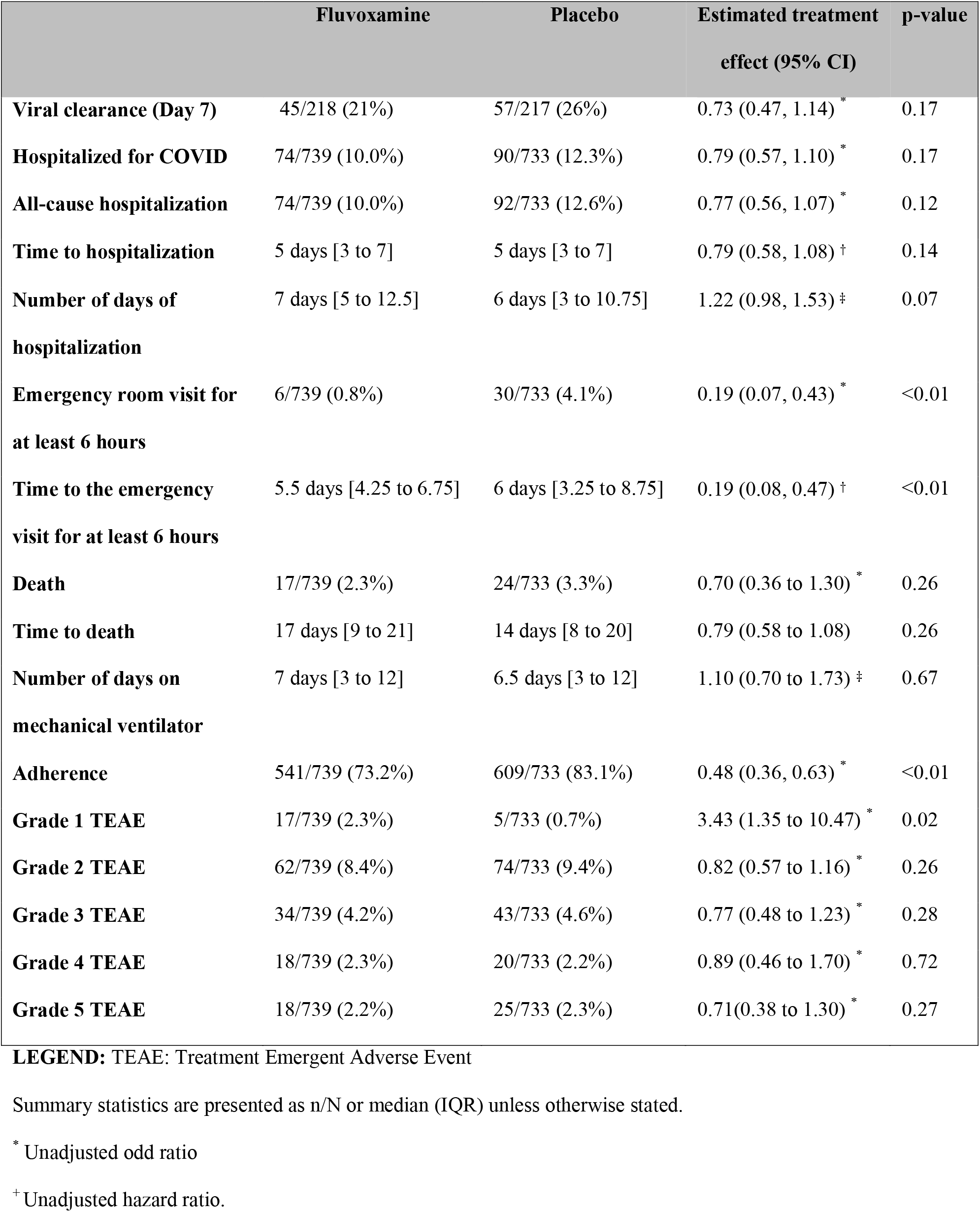
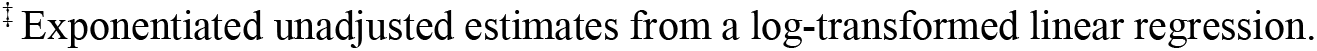
Secondary outcomes of fluvoxamine vs placebo in the TOGETHER Trial

|  | Fluvoxamine | Placebo | Estimated treatment<br>effect (95% CI) | p-value |
| --- | --- | --- | --- | --- |
| **Viral clearance (Day 7)** | 45/218 (21%) | 57/217 (26%) | 0.73 (0.47, 1.14) * | 0.17 |
| **Hospitalized for COVID** | 74/739 (10.0%) | 90/733 (12.3%) | 0.79 (0.57, 1.10) * | 0.17 |
| **All-cause hospitalization** | 74/739 (10.0%) | 92/733 (12.6%) | 0.77 (0.56, 1.07) * | 0.12 |
| **Time to hospitalization** | 5 days [3 to 7] | 5 days [3 to 7] | 0.79 (0.58, 1.08) † | 0.14 |
| **Number of days of**<br>**hospitalization** | 7 days [5 to 12.5] | 6 days [3 to 10.75] | 1.22 (0.98, 1.53) ‡ | 0.07 |
| **Emergency room visit for**<br>**at least 6 hours** | 6/739 (0.8%) | 30/733 (4.1%) | 0.19 (0.07, 0.43) * | <0.01 |
| **Time to the emergency**<br>**visit for at least 6 hours** | 5.5 days [4.25 to 6.75] | 6 days [3.25 to 8.75] | 0.19 (0.08, 0.47) † | <0.01 |
| **Death** | 17/739 (2.3%) | 24/733 (3.3%) | 0.70 (0.36 to 1.30) * | 0.26 |
| **Time to death** | 17 days [9 to 21] | 14 days [8 to 20] | 0.79 (0.58 to 1.08) | 0.26 |
| **Number of days on**<br>**mechanical ventilator** | 7 days [3 to 12] | 6.5 days [3 to 12] | 1.10 (0.70 to 1.73) ‡ | 0.67 |
| **Adherence** | 541/739 (73.2%) | 609/733 (83.1%) | 0.48 (0.36, 0.63) * | <0.01 |
| **Grade 1 TEAE** | 17/739 (2.3%) | 5/733 (0.7%) | 3.43 (1.35 to 10.47) * | 0.02 |
| **Grade 2 TEAE** | 62/739 (8.4%) | 74/733 (9.4%) | 0.82 (0.57 to 1.16) * | 0.26 |
| **Grade 3 TEAE** | 34/739 (4.2%) | 43/733 (4.6%) | 0.77 (0.48 to 1.23) * | 0.28 |
| **Grade 4 TEAE** | 18/739 (2.3%) | 20/733 (2.2%) | 0.89 (0.46 to 1.70) * | 0.72 |
| **Grade 5 TEAE** | 18/739 (2.2%) | 25/733 (2.3%) | 0.71(0.38 to 1.30) * | 0.27 |

**LEGEND:** TEAE: Treatment Emergent Adverse Event
Summary statistics are presented as n/N or median (IQR) unless otherwise stated.
\* Unadjusted odd ratio
† Unadjusted hazard ratio.
‡Exponentiated unadjusted estimates from a log-transformed linear regression.

### Sub-group Analyses

In the prespecified subgroup analysis, we found no evidence of moderation of treatment effect for fluvoxamine compared to placebo, for sub-groups of age, sex, days since symptom onset, or co-morbidities (Figure 3 and web-appendix eTable 1).

**Figure 3:**
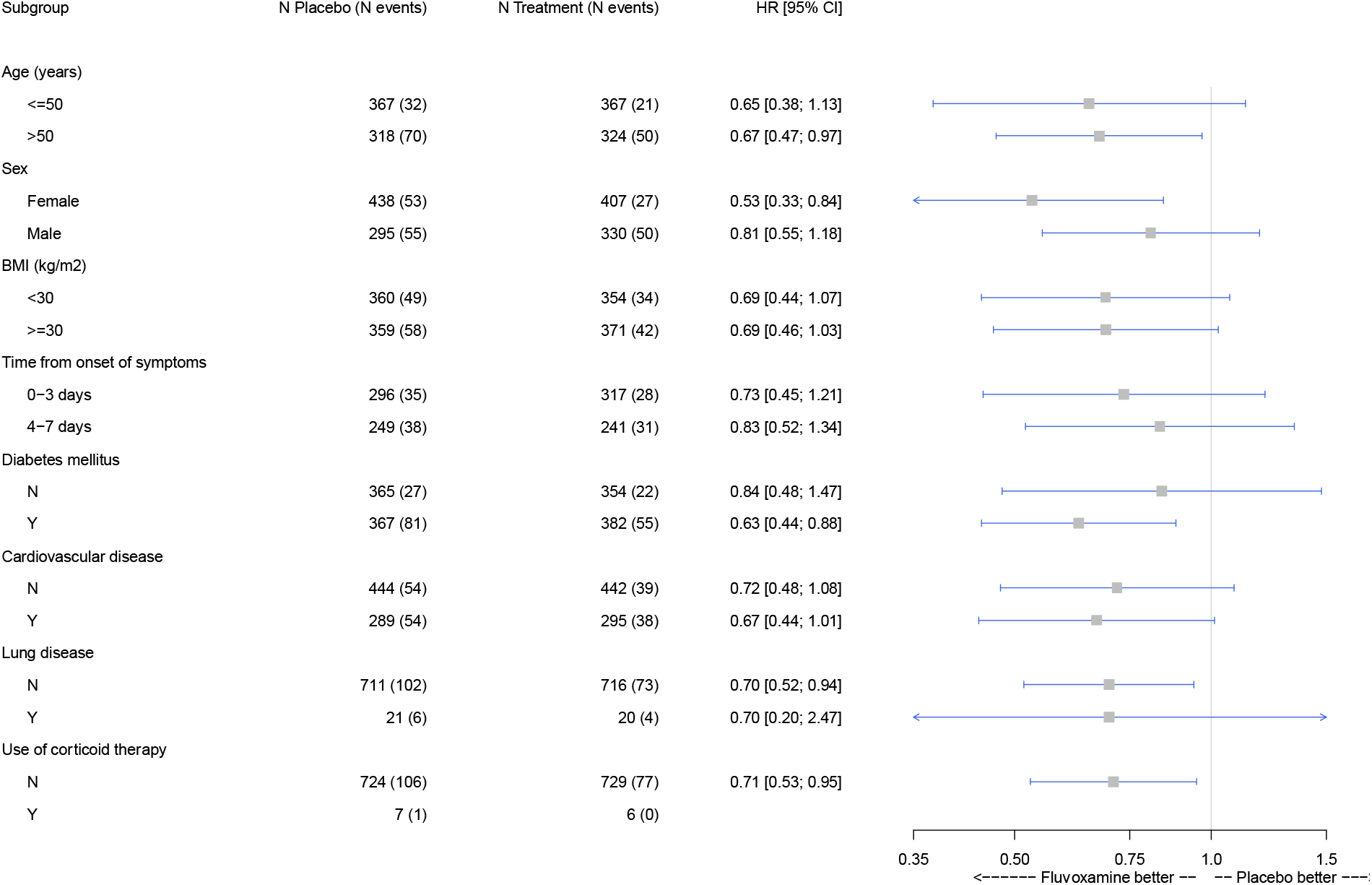
Sub-group analyses of fluvoxamine vs. placebo in the TOGETHER Trial

## DISCUSSION

This is the first large randomized controlled trial to test the efficacy of fluvoxamine for acute treatment of COVID-19. We found a clinically important absolute risk reduction of 4.3%, and 29% relative risk reduction, on the primary outcome of retention in an emergency setting for COVID-19 disease observation or hospitalization, consequent on the administration of fluvoxamine for 10 days. This study is only the 2^nd^ study to demonstrate an important treatment benefit for a repurposed drug in the early treatment population.^11^ Our findings represent the latest interim analysis of the trial resulting in the DSMC recommending stopping the active fluvoxamine arm. The final analysis from the trial, wherein all patients have contributed 28 days of follow-up data, will be made available 28 days after the last randomized patient has completed this period (August 25, 2021). Given fluvoxamine’s safety, tolerability, ease of use, low cost, and widespread availability, these findings may have influence on national and international guidelines on the clinical management of COVID-19.

### Comparison with prior evidence

Our results are consistent with an earlier smaller trial conducted in the United States (led by EJL and AMR).^2^ That study used a higher dose of fluvoxamine (100mg tid for 15 days) and included a lower risk group for the primary outcome but found no clinical deterioration among 80 patients receiving fluvoxamine vs. 6 of 72 patients receiving placebo. A large observational study from France involved a different population, 7230 hospitalized COVID-19 patients, and reported a reduction in use of intubation or death with use of SSRIs.^1^

The underlying mechanism of fluvoxamine for COVID-19 disease remains uncertain. Although hypotheses include several potential mechanisms,^6^ the main reason for the initial study of fluvoxamine as a treatment of COVID-19 was its anti-inflammatory action through activation of the σ-1 receptor (S1R).^12^ S1R is an endoplasmic reticulum (ER) chaperone membrane protein involved in many cellular functions,^13^ including regulation of ER stress response/unfolded protein response and regulation of cytokine production in response to inflammatory triggers.^14^ In the presence of fluvoxamine, S1R may prevent the ER stress sensor Inositol-Requiring Enzyme 1α (IRE1) from splicing and activating the mRNA of X-Box Protein 1 (XBP1), a key regulator of cytokine production including IL-6, IL-8, IL-1β and IL-12. In a 2019 study by Rosen and colleagues, fluvoxamine showed benefit in preclinical models of inflammation and sepsis through this mechanism.^14^

Another mechanism may be fluvoxamine’s anti-platelet activity.^15^ SSRIs can prevent loading of serotonin into platelets and inhibit platelet activation, that may reduce the risk of thrombosis, and these antiplatelet effects can be cardioprotective. In vitro and animal studies are needed to help clarify the most likely mechanism(s). Biomarker studies included as part of future RCTs may also help to clarify mechanisms.

### Strengths and limitations

Since the start of the COVID-19 pandemic, there have been more than 2800 RCTs registered on clinicaltrials.gov. However, less than 300 have been reported and the vast majority of clinical trials have been small and underpowered, with sample sizes less than 100. In many cases, these trials have been unsuccessful at recruiting as the local epidemics occur in waves and sustainable infrastructure to maintain staff or local interest for recruitment is lacking. The trials that provide the clearest medical understanding tend to be the larger platform trials, such as SOLIDARITY,^16^ RECOVERY,^17^ PRINCIPLE,^11^ and REMAP-CAP.^18^ As a result, we actively collaborate with other investigators running trials with overlapping interventions so that they can be aware of our study decisions and determine whether they should influence their respective trials.

Major strengths include the rapid recruitment and enrolment of high-risk patients for the development of severe COVID-19. Our recruitment strategy involves the engagement with the local public health system, thus allowing recruitment that frequently exceeds twenty patients per day. We enrolled only participants with diagnosed COVID-19 and less than 7 days of symptom onset using a commercially available COVID-19 AG rapid test (Panbio ®). The concordance of COVID-19 positive tests with RT-PCR was evaluated on the group of participants with PCR evaluations and found a concordance rate of > 99% on both tests collected at baseline.

Our understanding of the epidemiology of COVID-19 as well as its disease progression and outcomes have evolved since beginning this platform trial in June 2020. Early studies assessed the effects of interventions of viral load and clearance, while later studies also evaluate more clinical outcomes. We made adjustments to the trial according to prespecified rules and in communication with the appropriate ethics review committees that allowed us to respond to the epidemic waves while maintaining high rates of recruitment. Unlike many outpatient clinical trials, our study involves direct patient contact through the use of medical students, nurses and physicians who do at-home visits as well as follow-up via telecommunications. Given the rapid recruitment of patients in combination with the high event rate of emergency room visits and hospitalizations, we were able to evaluate the effects of interventions when portions of the planned population had been recruited. The period of time between first recruitment of a patient on fluvoxamine and the data cut for our trial was 197 days. Our trial assessed a primary outcome as a binary event having occurred by 28 days post-randomization.

One of the major limitations of our fluvoxamine trial is primarily related to the challenges of conducting a trial in a disease that is not well characterized. Currently, there is no standard of care that exists for early treatment of COVID-19 and various advocacy groups promote different interventions, including some of those evaluated in this and our previous trials. Furthermore, there is little understanding of who is at greatest risk of disease progression from this disease as some patients with numerous risk factors do recover quickly while some others with less established risk factors may not. Our population had a higher rate of hospitalization events than observed in most clinical trials, thus permitting inferences on treatment effects in this higher-risk population.

### Implications

Our trial has found that fluvoxamine, an inexpensive existing drug, reduces the need for advanced disease care in this high-risk population. A ten-day course of fluvoxamine costs approximately $4 even in well-resourced settings.^19^ Our study compares favorably and exceeds the treatment effects of more expensive treatments including monoclonal antibodies for outpatient treatment.^20,21^ The absolute number of serious adverse events associated with fluvoxamine was lower than our placebo group and this might reflect the modulatory effect of fluvoxamine on systemic inflammation in these participants. Lower respiratory tract infections were reported less frequently in patients in the fluvoxamine group than those in the placebo group. This is concordant with the reduction of hospital admissions in patients with confirmed COVID-19 treated with fluvoxamine, and the numerically lower number of patients requiring mechanical ventilation.

Fluvoxamine is widely available but is not on the WHO Essential Medicines List,^22^ whereas a closely related SSRI fluoxetine is on the list. It is now of critical importance to determine whether a class-effect exists and whether these drugs can be used interchangeably for COVID-19. The recent important findings that inhaled budesonide increased time to recovery among a similar population as our trial and had a trend towards decreased hospitalizations suggests this as an alternative or additional intervention for outpatient care that should be evaluated. The PRINCIPLE trial evaluated time to recovery using self-reported recovery up to 28 days after randomization to budesonide.^11^ Our trial differed as we evaluated improvement in the WHO categorization of disease disability up to days 14 and then 28 (web-appendix eFigure 2). Finally, our study was among unvaccinated patients. Further evidence of treatment benefits are needed to determine the effect of fluvoxamine among vaccinated populations.

Use of interventions, including fluvoxamine, to prevent progression of illness and hospitalization is critically dependent on identifying higher risk individuals. Unselected populations will have a lower risk. What absolute reduction in risk of clinical deterioration would motivate patients to choose treatment (probably the >4% we observed, but perhaps not much lower) remains uncertain. These considerations raise the importance of the development of a validated prediction rule for deterioration in patients in the early stages of COVID-19 infection.

### Conclusion

Administration of fluvoxamine reduced the rate of prolonged observation in an emergency care setting or hospitalization due to COVID-19 in people with a high risk of serious disease.

## Data Availability

Trial data are shared with the International COVID-19 Data Alliance (ICODA).

## ACKNOWLEGEMENTS

The trial was supported by FastGrants and the Rainwater Foundation. GR and EJM had full access to all the data in the study and take responsibility for the integrity of the data and the accuracy of the data analysis. Our research network consists of partnerships between academics and clinicians at McMaster University in Ontario, Canada, and Pontificia Universidade Catolica de Minas Gerais, Claros State University, and University of Ouro Preto in Minas Gerais, Brazil. Other partners include Cytel, Platform Life Sciences, MMS Holdings, WHO Therapeutic Guidelines Committee, and the Society for Clinical Trials. Trial documents are found on the Open Science Framework (https://doi.org/10.17605/OSF.IO/EG37X). Trial data are shared with the International COVID-19 Data Alliance (ICODA). The work has been presented and reviewed by the WHO Platform Trials Group.

## Data Safety and Monitoring Committee (DSMC) Members

William Cameron, University of Ottawa (Canada), James Orbinski, York University (Canada), Sonal Singh, University of Massachusetts (USA), Kristian Thorlund, McMaster University (Canada), Jonas Haggstrom of Cytel Inc. (Sweden).

