## Supplemental file 1 for "Effect of Early Treatment with Fluvoxamine on Risk of Emergency Care and Hospitalization Among Patients with COVID-19: The TOGETHER Randomized Platform Clinical Trial" (from another version of this preprint: version 1, 2021-08-23)

This appendix forms part of the original submission.

The COVID-19 TOGETHER initiative was designed to evaluate repurposed treatments for COVID-19 disease through an adaptive trial design in two arms being conducted in Brazil. The Brazilian TOGETHER arm was supported by a network of primary care research centers from the state of Minas Gerais, Brazil, devoted to a comprehensive evaluation and treatment of patient with COVID-19. The trial was fully integrated with local public health authorities (Brazilian Unified Health System – SUS) as part of coping strategy for COVID-19 pandemic. Namely, the main institutions involved were: Cardresearch – Cardiologia Assistencial e de Pesquisa. This initiative was funded by FastGrants and The Rainwater Foundation.

**Brazilian Executive Committee**

Gilmar Reis[1,2], Eduardo ASM Silva[1,2], Daniela CM Silva[1,2], Castilho VQ Santos[2], Ana MR Nogueira[3], Ana PFG Almeida[3], Adhemar DF Neto[4], Leonardo CM Savassi[5], Luciene B Ribeiro[1], Maria IC Simplicio[1], Thiago S Ferreira[1]

**Steering and Executive Committee**

Gilmar Reis[1,2], Lehana Thabane[6], Jamie Forrest[7,9,] Edward J Mills[6,7]

[1]Cardresearch – Cardiologia Assistencial e de Pesquisa, Belo Horizonte, Brazil; [2]Pontificia Universidade Catolica de Minas Gerais, Belo Horizonte; [3]Montes Claros State University, Montes Claros, Brazil; [4]Federal University of Juiz de Fora, Governador Valadares, Brazil; [5]Federal University of Ouro Preto, Ouro Preto, Brazil; [6]McMaster University, Hamilton, Canada, [7]Cytel Health, Vancouver, Canada, [9]University of British Columbia, Vancouver, Canada.

**Data Safety and Monitoring Committee**

William Cameron, University of Ottawa (Canada), James Orbinski, York University (Canada), Sonal Singh, University of Massachusetts (USA), Kristian Thorlund, McMaster University (Canada), Jonas Haggstrom of Cytel Inc. (Sweden).

**Clinical Events Classification Committee**

Gilmar Reis[1,2] (chair), Luciene B Ribeiro[1] (adjudicator), Thiago S Ferreira[1] (adjudicator), Ana MR Nogueira (adjudicator)[3], Laura C Oliveira[1] (operations manager), Heloísa A Costa[1] (operations manager)

[1]Cardresearch – Cardiologia Assistencial e de Pesquisa, Belo Horizonte, Brazil; [2]Pontificia Universidade Catolica de Minas Gerais, Belo Horizonte, Brazil;

**Trial Operations**

Maria IS Simplicio[1] (site manager), Thais C Siqueira[1] (site monitoring), Nayara S Drumont[1] (site monitoring), Leticia C. Siqueira[1] (site monitoring), Stephanny K Bessa[1] (site monitoring), João VB Vieira[1] (telemonitoring manager), Kenia SG Gonçalves[1] (site monitoring), Laura C Oliveira[1] (operations manager), Heloisa A Costa[1] (operations manager), Vania R Campos[1] (administrative manager), Aline LJ Martins (logistic manager), Rosemary M Oliveira[1] (international affairs manager), Carla SO França[1] (data management), Aline C Milagres[1] (data management), Luciene B Ribeiro[1] (regulatory affairs manager), Tainara S Vieira[1] (regulatory affairs), Lineria HM Suterio (pharmaceutical operations manager),

[1]Cardresearch – Cardiologia Assistencial e de Pesquisa, Belo Horizonte, Brazil;

**Participating Enrolling Centres:**
Included below are representatives from the enrolling centers at participating cities that enrolled at least 1 patient. Centres are listed in order of enrollment contribution. All study sites were located in the State of Minas Gerais, Brazil.

*Cardresearch – Cardiologia Assistencial e de Pesquisa, Belo Horizonte*

Aline Lúcia de Jesus Martins, Anna Clara de Jesus Oliveira, Bárbara Nogueira Roberti, Bruno Reis Garcia, Carina Daniela Lira Moreira Figueiredo, Carolina Couto de Azevedo Cysne, Clara Bensemann Gontijo Pereira, Ednilson Barbosa Dias, Emanuelle Cristina Soares Gonçalves, Fernanda Leal Guimarães, Flávio Eduardo Giorgeto, Gabriel Torga Saade Rodrigues, Júlia Campos de Rezende, Júlia Diniz Assis Moreira, Heloisa Alves Costa, João Victor Barbosa Silveira, Joselaine Gomes de Souza, Juliana Marques Santos Ferreira, Kênia Sthéfhane Guimarães Gonçalves, Laura Campos Oliveira, Letícia Campos Siqueira, Letícia de Fátimas Costa, Lorena Gabrielle Miranda Maciel, Lorena Giovanna Silva Peixoto, Lorena Mesquita Alves, Luana Rosa Rodrigues, Luciene Barra Ribeiro, Luisa Andrade de Almeida, Marcela Luíza Amaral Resende Lara, Maria Izabel Campos Simplicio, Maria Luiza Magalhães Parreiras, Mariana Campos Siqueira, Mariana Cristina Flores Mariano, Marina Guimarães Bragatto, Matheus Andrade Almeida e Silva, Mayra Estefane Silva dos Santos, Nayara Santos Drumond, Pollyana Maria Silva Diniz, Roberta Dumont Paiva Lopes, Rosemary Muniz de Oliveira, Stephanny Kelvyn Bessa, Tainara Silva Vieira, Tatiana Aparecida Gomes da Silva, Thiago Santiago Ferreira, Túlio Fonseca e Silva Quadros, Thais Campos Siqueira, Vânia Regina Carmo, Vitor Augusto Lima do Vale, Vitor Vilaça Oliveira, ,

*City of Ibirité*

Aline Cruz Milagres, Ana Carolina Aguiar Estevam, Ana Martins de Oliveira, André Henrique de Sousa Oliveira, Carla Stefany Oliveira França, Laylla Michelle Pereira, Luciléia Alcântara Gomes e Silva, Patrícia Santana Assis, Regiane Aparecida de Andrade, Thaís de Paula Ferreira da Silva, Thiago de Alcântara César

*City of Sete Lagoas*

Amanda Luiza do Espírito Santo Pinheiro, Ana Carolina Matos Ferreira, Ana Caroline Santos Silva, Ana Luiza Silva Taveira, Bruno Victor de Souza Lima, Castilho Vitor Quirino dos Santos, Débora Siqueira Diana, Emanuela Pontes Pereira Silveira, Francisca Lina Campos

Gonçalves, Gabriel Corrêa Costa, Geraldo César Barroso de Souza, Guilherme Aurelio D'Oliveira Alves, Isadora Luiza Teixeira Neto, Jordanna Porto Inácio, Lara Silveira Marques, Larissa Martins do Nascimento, Larissa Oliveira Moreira, Lorena Luiza Soares de Oliveira, Lucas Eduardo Lessa Mussi, Lucas Medeiros Ruas, Luiza Nogueira Assis Barbosa, Marcelo Rodrigues da Costa Fernandes, Maria Eduarda Guimarães de Souza, Maria Thereza Figueiredo Bélem Calazans, Mariana Rodrigues Marinho de Bastos, Pedro Henrique Arcanjo Alvarenga, Quimberli Vassinave Cujuí, Vinícius Alves Corrêa, Vitória Aparecida Cunha, Viviane Costa Santos

*City of Betim*

Ana Flávia Gomes Viana, Daniel Domingues Barbosa, Daniela Carla Medeiros, Eduardo Augusto dos Santos Moreira Silva, Felipe Menezes Andrade, Gabriela Ferreira Reis, Giovane Carvalho Constantino Ferreira de Paula, Jéssica Aguilar Silva, Lucas Júnio da Silva, Luiz Gustavo Fernandes Dias, Marcela Luiza Amaral Resende Lara, Maria Luisa Lara Lanza Stabile

*City of Santa Luzia*

Bernardo Saraiva Resende de Camargos, Camila Araújo Heringer, Daniel Jacinto Mendonça Filho, Diuly Ane Faria Rezende, Higor Gomes Mussi, Igor Bernardes Caciquinho, João Pedro Ribeiro Santos,João Ricardo Garcia Costa, José Lauro Guimarães Cardoso, Lorena Amaral de Oliveira, Luana Rocha Oliveira Matos, Mariana Cavaliere Batista e Silva, Matheus Lucca Ângelo Costa Rodrigues, Mateus de Oliveira Ferreira, Murilo Passinati Perez, Natalia Batista Zanetti, Sarah Jeniffer da Silva Jamar, Sofia Caporalli Barbosa, Sofia Vianna Von Bentzeen Rios, Stella Silva Rosa, Thomaz Takashi Ferreira Morimoto, Vitória Helena de Souza Campos, Vitor Augusto Alves da Silva

*City of Nova Lima*

Alice Pinheiro Vanetti, Ana Carolina Araújo Lage Santos, Felipe Viana Santos, Fernanda Freitas Pinheiro, Fernanda Perez Rocha, Isabela Gontijo Mendonça, Isadora Gomides Faria, Larissa Milagres Mol, Letícia de Fátima Costa, Suzana Cristina Ricco, Thiago Araújo do Nascimento

*UniFIPMoc - City of Montes Claros,*

Ana Maria Ribeiro Nogueira, Ana Paula Figueiredo Guimarães de Almeida, Ana Paula Guimarães Alves de Carvalho, Antônio Henrique Batista Jorge, Artur Pimenta Ribeiro, Cleide Rocha Veloso, Elisângela Santos Sema, Gonçalvino Eleutério Murta, João Warley Alves, Jorge Fernando Rocha Veloso, Joyce Souto Barbosa Pimenta de Figueiredo, Karolina Campos Sampaio Lopes, Loren Montanha Costa, Ludimila Pereira de Souza, Maria de Fátima César Lima, Maria Izabel Silveira Gonçalves, Marizete Gislaine Mendes, Mayra Darlliane Loiola Silva, Samuel de Paiva Oliveira, Thiago Rodrigues Ferro

*City of Brumadinho*

Ana Beatriz Amorim, Carolina Francisca Martins, Eduardo Diniz Callegari, Huisnei Ferreira Lourenço, Jussara Carolina Assis Ribeiro Nascimento, Solange Santos Moreira Sales

*City of Governador Valadares*

Adhemar Dias de Figueiredo Neto, Adriana Marcos Mendes Rabelo, Aline do Carmo Rosa , Ana Paula Vilas Boas Wherberth, Arthur Henrique Nunes Leite Oliveira, André Gustavo Pinto de Souza, Damiana Rogai Siqueira, Felipe Coelho Soares de Oliveira, Felipe Fraga Damaceno, Guilherme Rhis, Iandra Silva Almeida, Igor Barros de Araújo Borges, Igor Brandão Rocha, Jorge Carlos do Amaral Júnior, José Pio Marques II, Laura Lima Vargas, Luiz Eduardo Coelho Fava, Luiza de Azevedo Freitas Giles , Marina Lacerda Marques, Maura Alina Morais Veríssimo, Neila Rodrigues Vargas de Paula, Pâmela de Sousa Dias Demuner, Rafaella Rosa de Oliveira Fernandes, Ramail Santos Pouzas, Roberta Coelho de Marco, Ronan Figueiredo Mourão, Sabrina Stefany da Silva Souza, Sara Silva Ferraz

*Federal University of Ouro Preto, City of Ouro Preto*

Aline Dias Bedetti, Anna Carolina Motta Costa, Cristiana Silva Mascarenhas, Diana Luiza Marinho Brandão, Felipe César Soares, Flávia Marcella Sena Gonçalves Borba,
Karinna Guimarães Gomes, Leonardo Cançado Monteiro Savassi, Marina Eduarda Santos, Maryane de Oliveira Silva, Rayane Elen Fernandes Silva, Ruan Carling Schott Wondollinger, Yuri Barbosa de Menezes

*Santa Casa de Misericórdia, City of Sabará*

Bárbara Ellen Souza Rezende, Graziela César de Sousa, Graziele Aparecida Silva, Isabela Cristina de Oliveira Campos, Lara Silva Souza, Laura Nacife Rabello, Mariana Cristina Flores Emiliano, Roberta Ellen Santos Oliveira

*Public health authorities and mayors*
We are in debt with the following local public health authorities and mayors (listed by enrollment):

**City of Ibirité**
William Parreira Duarte (Mayor), Carina Bitarães (public health authority)

**City of Sete Lagoas**
Duílio de Castro (Mayor), Flávio Pimenta Silveira (public health authority), Alber Alípio Ribeiro (public health authority)

**City of Betim**
Vittorio Medioli (Mayor), Augusto Viana da Rocha (public health authority), Hilton Soares de Oliveira (public health authority)

**City of Santa Luzia**
Christiano Augusto Xavier Pereira (Mayor) Nádia Cristina Dias Duarte Tomé (public health authority)

**City of Nova Lima**
João Marcelo Diegues Pereira (Mayor), Diogo Jonata Ribeiro (public health authority)

**City of Montes Claros**
Humberto Guimarães Souto (Mayor), Dulce Pimenta Gonçalves (public health authority)

**City of Brumadinho**
Avimar de Melo Barcelos (Mayor), Eduardo Diniz Callegari (public health authority)

**City of Governador Valadares**
André Luiz Coelho Merlo (Mayor), Edna Gomes Leite (public health authority)

**City of Ouro Preto**
Angelo Oswaldo de Araujo Santos (Mayor), Glauciane Resende do Nascimento (public health authority)

**Supplementary Tables and Figures**

**eFigure 1: Time to hospitalization or extended emergency room visit due to COVID-19**

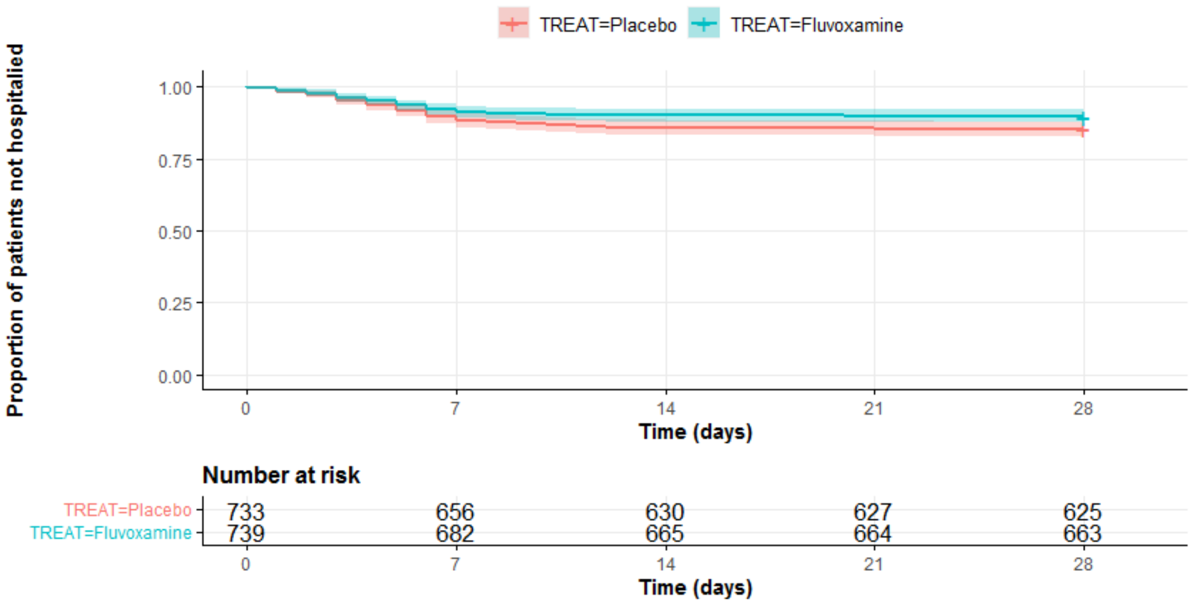

### eFigure 2: Time to symptom resolution (WHO symptom index scale)

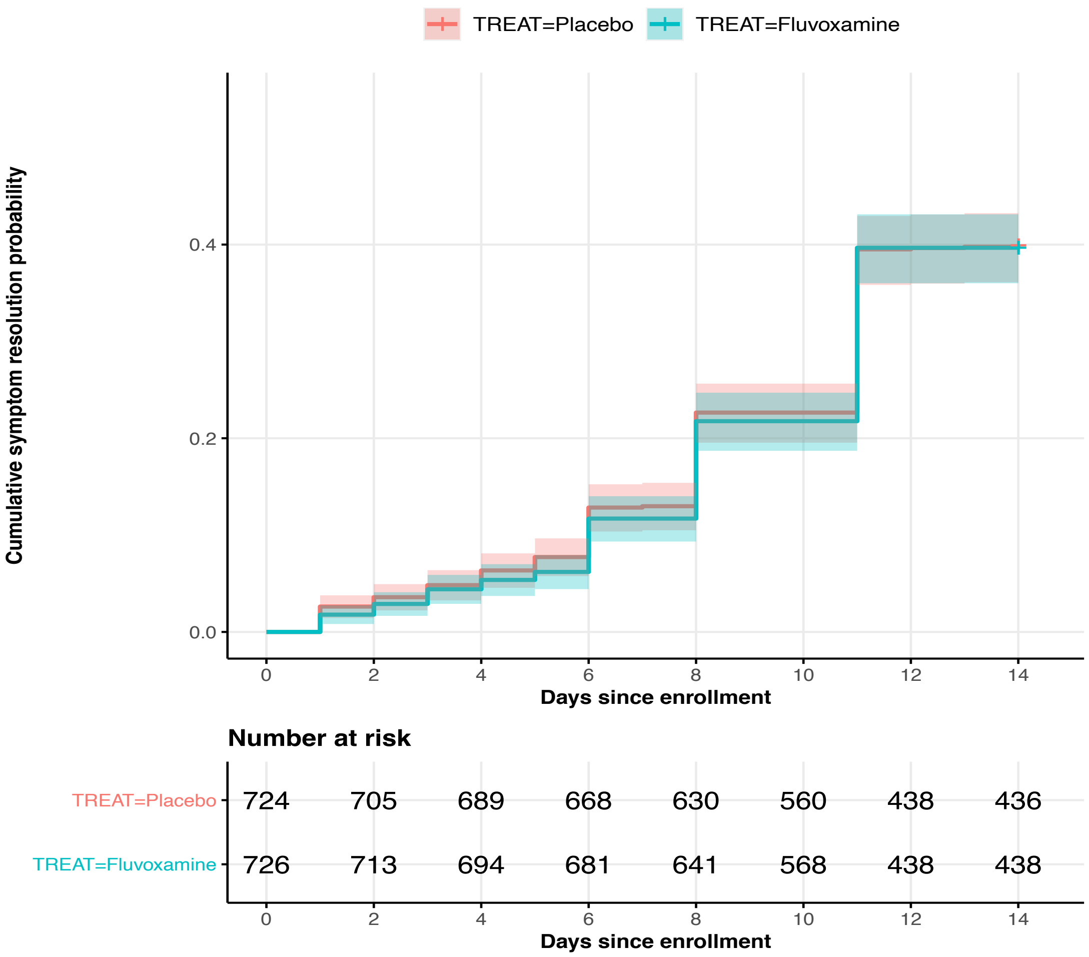

### eFigure3: Probability of viral detection (Days 0, 3, 7)

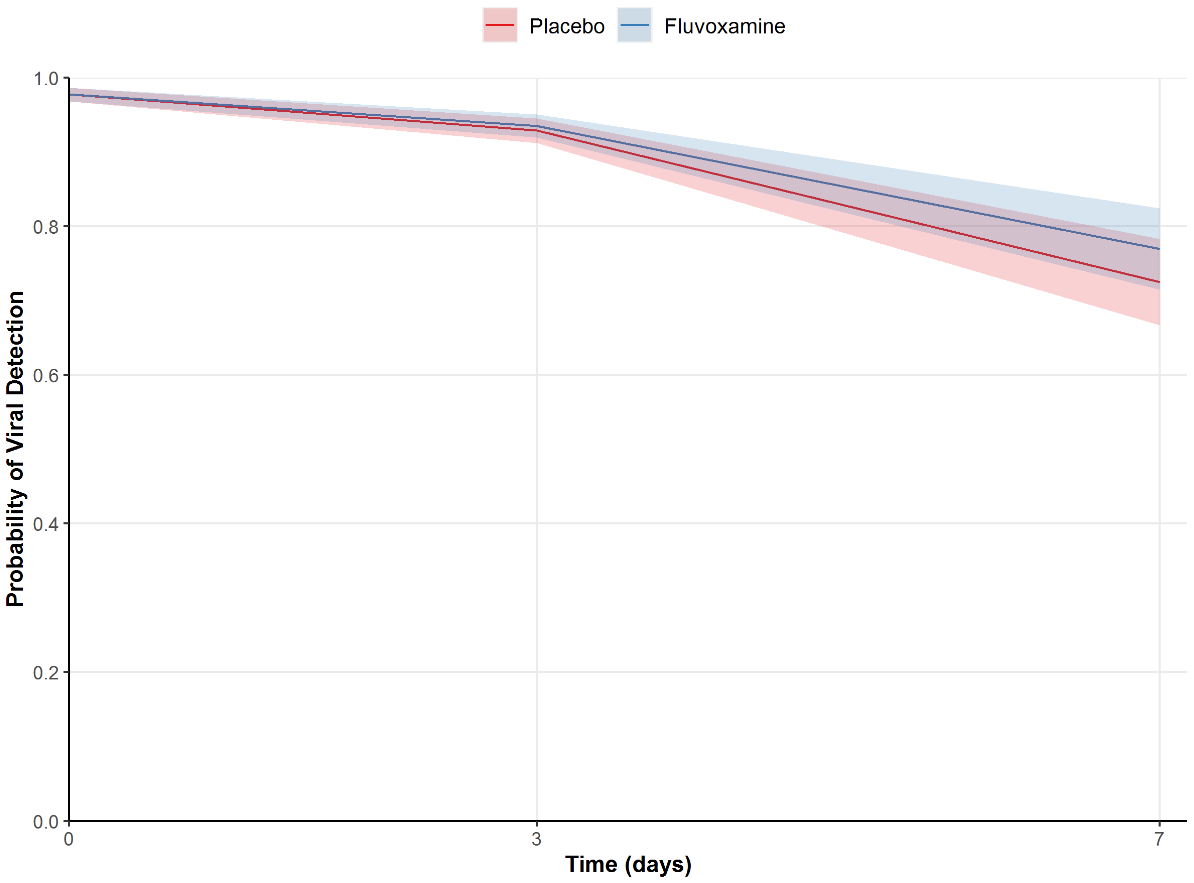

**eTable1: Summary of Sub-Group Analysis for Hospitalization or Extended Emergency Room Visit Due to COVID-19**

| Factor | Subgroup | N_Placebo | N_Fluvoxamine | n_Placebo | n_Fluvoxamine | HR (95% CI) |
|---|---|---|---|---|---|---|
| Age | <=50 years | 367 | 367 | 32 | 21 | 0.65 [0.37; 1.12] |
| Age | >50 years | 318 | 324 | 70 | 50 | 0.69 [0.48; 0.99] |
| Sex | Female | 438 | 407 | 53 | 27 | 0.53 [0.34; 0.85] |
| Sex | Male | 295 | 330 | 55 | 50 | 0.82 [0.56; 1.20] |
| Time from onset of symptoms | 0-3 days | 296 | 317 | 35 | 28 | 0.74 [0.45; 1.22] |
| Time from onset of symptoms | 4-7 days | 249 | 241 | 38 | 31 | 0.83 [0.52; 1.34] |
| Diabetes mellitus | N | 365 | 354 | 27 | 22 | 0.83 [0.47; 1.46] |
| Diabetes mellitus | Y | 367 | 382 | 81 | 55 | 0.64 [0.46; 0.90] |
| Cardiovascular disease | N | 444 | 442 | 54 | 39 | 0.71 [0.47; 1.08] |
| Cardiovascular disease | Y | 289 | 295 | 54 | 38 | 0.68 [0.45; 1.04] |
| Lung disease | N | 711 | 716 | 102 | 73 | 0.70 [0.52; 0.95] |
| Lung disease | Y | 21 | 20 | 6 | 4 | 0.69 [0.19; 2.45] |
| Use of corticoid therapy | N | 724 | 729 | 106 | 77 | 0.71 [0.53; 0.96] |
| Use of corticoid therapy | Y | 7 | 6 | 1 | 0 | NA |

**N, number of patients; NA, not applicable; HR, hazard ratio; CI, confidence interval.**
